# Potentially harmful consequences of artificial intelligence (AI) chatbot use among patients with mental illness: Early data from a large psychiatric service system

**DOI:** 10.1101/2025.11.19.25340580

**Authors:** Sidse Godske Olsen, Christian Jon Reinecke-Tellefsen, Søren Dinesen Østergaard

**Affiliations:** Department of Affective Disorders, Aarhus University Hospital – Psychiatry, Aarhus, Denmark; Department of Clinical Medicine, Aarhus University, Aarhus, Denmark

**Keywords:** Generative Artificial Intelligence, Delusions, Mania, Obsessive-Compulsive Disorder, Feeding and Eating Disorders, Suicidal Ideation

## Abstract

Chatbots driven by generative artificial intelligence (AI chatbots) have become ubiquitous. Recently, concerns have risen over the possibility that use of AI chatbots may be harmful for people prone to mental illness. Here, we aimed to investigate whether there are reports compatible with potentially harmful consequences of AI chatbot use on mental health among patients with mental illness receiving care in a large psychiatric service system. Based on screening of clinical notes from electronic health records mentioning ‘chatbot’ or ‘ChatGPT’ from the Psychiatric Services of the Central Denmark Region, we report on 38 patients for whom use of AI chatbots had potentially harmful consequences for their mental health - with worsening/consolidation of delusions being the most common presentation. Mental health professionals should be aware of this possibility and guide their patients accordingly, as it seems that some patients would likely benefit from reduced/no use of AI chatbots in their current form.

---

Chatbots driven by generative artificial intelligence (AI chatbots) have become ubiquitous.^1^ While the large language model technology underlying these tools may have a huge potential for societies at large, concerns—and substantial anecdotal evidence^2^— have risen over the possibility that use of AI chatbots may be harmful for people prone to mental illness.^3-5^ Specifically, it seems that interaction with AI chatbots, especially if intense/of long duration, may contribute to onset or worsening of delusions or mania – with severe or even fatal consequences.^3-5^ Given the large uptake of this technology, ChatGPT—the clear market leader—passed 900 million downloads in July 2025,^1^ this could pose a tangible threat to public mental health. At this stage, however, almost all reports on potentially harmful consequences of AI chatbots stems from news media or online fora^2^ – and should be interpreted with the inherent limitations of these outlets in mind. Conversely, to our knowledge, there are very few accounts of this phenomenon from psychiatric services. Therefore, we aimed to investigate whether there are reports compatible with potentially harmful consequences of AI chatbot use on mental health among patients with mental illness receiving care in a large psychiatric service system.

This study was conducted in accordance with the method used to assess the potential impact of the COVID-19 pandemic^6^ and the 2022 Russian invasion of Ukraine^7^ on the mental health of patients with mental illness. Specifically, we identified all patients registered with at least one contact with the Psychiatric Services of the Central Denmark Region (CDR)—one of five Danish regions providing inpatient, outpatient and emergency psychiatric care to its approximately 1.4 million inhabitants—in the period from 1. September 1, 2022, to June 12, 2025 (ChatGPT was launched publicly on November 30, 2022). Subsequently, we searched all clinical notes in the electronic health records of these patients for the words “chatbot” and “ChatGPT” (not case sensitive) along with the following 10 alternative spellings/misspellings for each: “chat bot”, “chat-bot”, “chattbot”, “chatboot”, “chatbott”, “chatbotts”, “chatbote”, “chatbox”, “chabot”, “chatpot”, “Chat GTP”, “ChatGBT”, “ChatGRT”, “ChatGPTT”, “Chat GPT”, “Chat-GPT”, “ChattGPT”, “ChatJPT”, “ChatGBT”, and “ChatGPT3”. These 20 alternatives were provided by ChatGPT based on the following prompt: “Please create a list of the top 20 likely misspellings or variations of the words *ChatGPT* and *chatbot* that might arise from common human spelling errors, typos, or misunderstandings of the correct terms. Provide 10 variations for each word”. We chose to include only ChatGPT and no other AI chatbot names as search term due to the uptake dominance of ChatGPT^1^ and our impression of genericization of this trademark (including in Denmark).

The notes identified by this search were assessed independently by SGO and CJR-T to determine whether any of the identified notes was compatible with potentially harmful consequences of use of AI chatbots on mental health (cases of doubt/discrepancy were discussed until consensus was reached). Finally, the cases of potentially harmful consequences were labelled according to the dominant type of psychopathology in agreement with Rohde et al.^6^ (see the Supplementary Material for details). This was first done independently by SGO and CJR-T and subsequently cases of doubt/discrepancy were discussed until consensus was reached).

The study was approved by the Legal Office of the CDR, which also waived the need for obtaining patient consent in agreement with the Danish Health Care Act, §46, Section 2 (Approval no. 1-45-70-58-25). Studies based solely on electronic health record data are exempt from ethical review board approval in Denmark (Waiver no. 1-10-72-116-25).

We found that a total of 53,974 patients (52% females, median age 27 years (25%-75%: 17-44 years)) had at least one contact with the Psychiatric Services of the CDR in the period from September 1, 2022, to June 12, 2025. During this period, 10,712,856 notes were entered into the electronic health record system. Among these, 181 notes from 126 unique patients (51% females, median age 28 years (25%-75%: 21-37 years)) contained at least one of the 22 search terms with an increased rate over time (see Figure 1).

**Figure 1.**
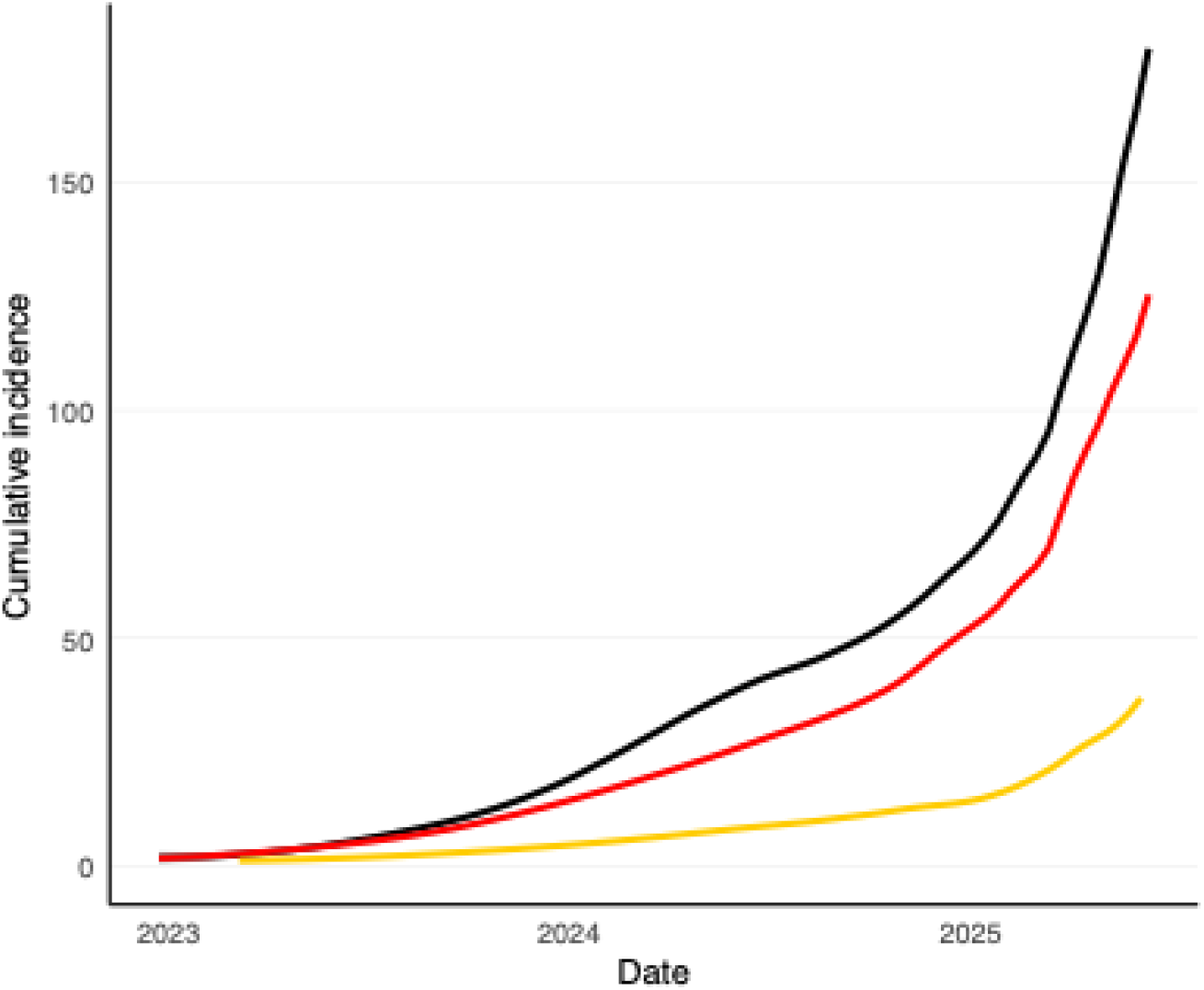
Cumulative incidence over time of 1) clinical notes containing one of the 22 chatbot/ChatGPT search terms, 2) unique patients with at least one of these clinical notes, and 3) unique patients with a clinical note compatible with potentially harmful consequences of use of AI chatbots on mental health Locally weighted scatterplot smoothing was applied to the cumulative incidence curves with an alpha of 0.75

The result of the consensus assessment was that among the 181 notes containing one of the 22 chatbot/ChatGPT search terms, notes from 38 unique patients (39% females, median age 28 years, 25%-75%: 22-39 years) were compatible with potentially harmful consequences of use of AI chatbots on mental health. Due to risk of identification, we are not allowed to describe the exact psychopathology of the 38 cases, but it belonged to the following overarching categories (see the Supplementary Material) ordered by cumulative incidence (when n<5, numbers are not reported due to risk of identification): Delusions (n=11), suicidality/self-harm (n=6), feeding or eating disorder (n=5), mania/hypomania/mixed state (n<5), obsessions or compulsions (n<5), depression (n<5), anxiety (n<5), other symptoms/miscellaneous n<5), ADHD-related symptoms(n<5), and unspecific stress n<5). Notably, the descriptions in the notes in question were thematically compatible with descriptions reported elsewhere, e.g., AI chatbots as object/consolidator of delusions, ^2^AI chatbots reinforcing hypomania/mania,^4^ AI chatbots being used compulsively in an attempt to relieve obsessions,^8^ AI chatbots enabling calorie restriction/counting,^9^ and AI chatbots being used to seek information on suicide methods.^10^

There were also examples of patients using AI chatbots for either “talk therapy”/companionship against loneliness or diagnostics (e.g., entering symptoms and requesting an interpretation) despite these tools generally not having been developed nor validated for these purposes, and the legal liability of the companies behind the AI chatbots in cases of their products providing wrong/harmful advice being unclear. Finally, there were descriptions of more constructive use of AI chatbots (aiding in various practical tasks unlikely to cause harm).

To our knowledge, this is the first indication of potentially harmful consequences of AI chatbot use on mental health among patients with mental illness stemming from a study based on data from a large psychiatric service system. The results must, however, be interpreted in the light of the following limitations. First and foremost, the descriptions in the clinical notes are, by no means, evidence of a causal effect (e.g., there is no knowledge of the counterfactual: i.e., what would have happened had the patients not interacted with an AI chatbot). Second, this study is based on data from everyday clinical practice where the patients were not systematically questioned about AI chatbot use. Third, we employed a quite narrow search focusing exclusively on 22 search terms (chatbot, ChatGPT and 10 alternative spellings of each of the two). It follows from the two latter limitations that our results should also not be interpreted from an absolute perspective, i.e. the results do not speak to the incidence rate of potentially harmful consequences of AI chatbot use among patients with mental illness.

In conclusion, with the substantial caveats described above in mind, the results of this study support the notion that use of AI chatbots may have a negative impact on the mental health of patients with mental illness, especially regarding delusions. Mental health professionals should be aware of this possibility and guide their patients accordingly, as it seems that some patients would likely benefit from reduced/no use of AI chatbots in their current form.

## Acknowledgements

The authors are grateful to Bettina Nørremark and Anders Ørberg from the Psychiatric Services of the Central Denmark Region for their assistance with extraction and visualization of data.

## Data availability statement

The data cannot be shared due to restrictions enforced by Danish law for protecting patient privacy.

## REFERENCES

1. Storyboard 18. ChatGPT surpasses 900 million downloads, widens lead over rival AI chatbots. https://www.storyboard18.com/digital/chatgpt-surpasses-900-million-downloads-widens-lead-over-rival-ai-chatbots-75192.htm

2. Morrin H, Nicholls L, Levin M, et al. Delusions by design? How everyday AIs might be fuelling psychosis (and what can be done about it) PsyArXiv Preprints. 2025;doi:10.31234/osf.io/cmy7n_v5

3. Østergaard SD. Will Generative Artificial Intelligence Chatbots Generate Delusions in Individuals Prone to Psychosis? Schizophr Bull. Nov 29 2023;49(6):1418–1419. doi:10.1093/schbul/sbad128

4. Østergaard SD. Emotion contagion through interaction with generative artificial intelligence chatbots may contribute to development and maintenance of mania. Acta Neuropsychiatr. Aug 22 2025:1–9. doi:10.1017/neu.2025.10035

5. Østergaard SD. Generative Artificial Intelligence Chatbots and Delusions: From Guesswork to Emerging Cases. Acta Psychiatr Scand. Aug 5 2025;doi:10.1111/acps.70022

6. Rohde C, Hougaard Jefsen O, Nørremark B, Aalkjær Danielsen A, Østergaard SD. Psychiatric Symptoms Related to the COVID-19 Pandemic. Acta Neuropsychiatr. May 21 2020:1–7. doi:10.1017/neu.2020.24

7. Østergaard SD, Rohde C, Jefsen OH. Deterioration of patients with mental disorders in Denmark coinciding with the invasion of Ukraine. Acta Psychiatr Scand. Aug 2022;146(2):107–109. doi:10.1111/acps.13440

8. Vox. ChatGPT and OCD are a dangerous combo. https://www.vox.com/future-perfect/417644/ai-chatgpt-ocd-obsessive-compulsive-disorder-chatbots

9. Psychiatrist.com. NEDA Suspends AI Chatbot for Giving Harmful Eating Disorder Advice. https://www.psychiatrist.com/news/neda-suspends-ai-chatbot-for-giving-harmful-eating-disorder-advice/

10. The Guardian. ChatGPT encouraged Adam Raine’s suicidal thoughts. His family’s lawyer says OpenAI knew it was broken. 2025. https://www.theguardian.com/us-news/2025/aug/29/chatgpt-suicide-openai-sam-altman-adam-raine

