## Supplementary Material for "Potentially harmful consequences of artificial intelligence (AI) chatbot use among patients with mental illness: Early data from a large psychiatric service system"

**Labeling of psychopathology**

All records describing potentially harmful use of AI chatbots were labeled based on the following code:

**1. Substance abuse**

**2. Delusions**

**3. Hallucinations**

**4. Negative symptoms**

**5. Mania/hypomania/mixed state (**”irritable”, ”restless”, overconfident or risky behavior etc.)

**6. Depression** (”sad”, ”unhappy”, ”lonely”, ”down”, ”hopeless”, ”guilty” etc.)

**7. Suicidality/self-harm**

**8. Anxiety (**”worried”, ”anxious”, ”tense”, ”frightened”, ”panicky”, + autonomic symptoms)

**9. Trauma- or stressor-related symptoms** (”alert”, ”vigilant”, “flashbacks”, ”nightmares”, etc.)

**10. Obsessions or compulsions**

**11. Feeding or eating disorder** (anorexia, bulimia, binge eating)

**12. Autism spectrum symptoms** (exacerbated internalizing or externalizing behaviors)

**13. ADHD-related symptoms** (impulsivity, hyperactivity, attention difficulties)

**14. Disruptive behaviour in children** (antisocial, defiant, oppositional, violent)

**15. Tics (motor/verbal)**

**16. Aggression** (“anger”, “frustration”, “externalizing behaviors”)

**17. Unspecific stress** (”stressed”, ”under pressure”, ”dissatisfaction”/”decreased well-being**”**) not better accounted for by the above symptom domains.

**18. Other symptoms / miscellaneous**

Only one label (the dominant psychopathology) was given per clinical note.

The categories stem from: Rohde C, Hougaard Jefsen O, Nørremark B, Aalkjær Danielsen A, Østergaard SD. Psychiatric Symptoms Related to the COVID-19 Pandemic. *Acta Neuropsychiatr*. May 21 2020:1-7. doi:10.1017/neu.2020.24
